# Vitamin D and acute respiratory infection: secondary analysis of a previous randomised controlled trial and updated meta-analyses

**DOI:** 10.1101/2022.02.03.22270409

**Authors:** Mark J Bolland, Alison Avenell, Andrew Grey, Greg Gamble

**Affiliations:** Department of Medicine, University of Auckland, Auckland, New Zealand; Health Services Research Unit, University of Aberdeen, Foresterhill, Aberdeen, Scotland

**Keywords:** Vitamin D supplementation, Respiratory tract infections, Covid-19, Cluster randomised trials

## Abstract

**Background:** Recent meta-analyses concluded that vitamin D supplementation can prevent acute respiratory infection (ARI). However, the findings were heavily influenced by results from two arms of a six-arm cluster-randomised trial that were analysed without accounting for the cluster trial design. We have used publicly available data to provide results from the remaining four unpublished trial arms and to reanalyse the meta-analyses, accounting for the cluster trial design.

**Methods:** The intracluster correlation co-efficient (ICC) and design effect were estimated. We then calculated the risk reduction (RR) of ARI from summary statistics, adjusting for the design effect, individually for the five different vitamin D treatment groups (four previously unpublished) and for all the vitamin D groups pooled. For this trial, individual patient data were used to estimate the effect of vitamin D on ARI risk and number of ARIs, adjusting for the cluster trial design, using random-effects models. Finally, we reanalysed the most recent trial-level meta-analysis, including the trial data generated by the correct analysis of the cluster randomized trial.

**Results:** There were 744 trial participants (6 treatment groups, 21 clusters, mean cluster size 35.4). The ICC was 0.08 (95% CI 0.02-0.14) and design effect 3.75. In analyses based on summary statistics, there was no statistically significant effect of vitamin D on ARI risk in any individual treatment group, or when groups were pooled (RR 0.75, 95%CI 0.50-1.13). In individual patient data analyses, there was also no statistically significant effect of vitamin D on the ARI risk or number of ARIs in any treatment group, or when pooled: odds ratio 0.58 (0.26-1.29), rate ratio 0.70 (0.44-1.12), respectively. Update of the previous meta-analysis showed no effect of vitamin D on ARI either when data from the two arms of the trial, or when all trial arms were incorporated (RR 0.98, 0.96-1.00, P=0.10 both analyses).

**Conclusions:** Overall, vitamin D supplementation had no effect on the risk of an ARI or on the number of ARIs in this trial or in a re-analysis of the most recent meta-analysis. The results of the updated meta-analysis do not suggest that vitamin D supplementation would reduce the risk of Covid 19.

## Introduction

The possible effect of vitamin D supplementation on acute respiratory infection (ARI) has been the subject of a great deal of recent commentary, particularly in the context of Covid-19. Two recent high profile meta-analyses from the same research group have concluded that vitamin D supplements can reduce the risk of ARI [1, 2]. The results of both meta-analyses are heavily influenced by the Blue Sky study, a cluster-randomised trial of vitamin D in schoolchildren from Mongolia [3]. Data on ARI from two of the six groups of the trial (groups allocated to Mongolian milk with or without vitamin D) were published in 2012 [4], but ARI data from the other four vitamin D arms remain unpublished. One example of the influence of this trial report is that in the first meta-analysis (using individual patient data), daily or weekly vitamin D reduced the odds of ARI by 70% in the subgroup with baseline 25-hydroxyvitamin D (25OHD) <25 nmol/L [1]. Of the 234 individuals in this analysis, 192 (92%) were from the Blue Sky study. Another example is that if the trial is excluded from the second meta-analysis (which evaluated trial-level summary data) [2], the heterogeneity between the individual trial results decreases and the overall result changes from a statistically significant to a non-significant result (37 studies, odds ratio 0.92, 0.86-0.99, I^2^= 36% to 36 studies, 0.94, 0.88-1.01, I^2^= 25%). Additionally, in both meta-analyses the Blue Sky trial data were analysed as though the trial was individually randomised rather than cluster-randomised. This approach is incorrect as it over-estimates the precision of the effect size [5, 6]. The recommended approach is to adjust the results of a cluster-randomised trial for the clustering of participants inherent in the design. Such adjustments can be done on summary statistics or in regression analyses using individual patient data.

Given the interest in the topic and the high frequency of vitamin D supplementation trials, it seems likely further meta-analyses will be conducted. Rich-Edwards and colleagues have published a partial dataset from the Blue Sky trial (trial registration NCT00886379) [7]. We used this publicly available dataset to provide more data about the trial results, including the intracluster correlation coefficient (ICC) and design effect, to allow summary data from the trial to be correctly incorporated in future meta-analyses, and to report results from the four previously unpublished vitamin D supplementation groups in the trial. We then updated the most recent trial-level meta-analysis [2] using these data, analysed appropriately for their cluster design.

## Methods

The trial details have been described previously [3, 4]. Briefly, this was a cluster-randomised trial in which 21 classrooms of 9 to 11-year-old schoolchildren from Mongolia were randomized to 6 different groups; five treatment groups which each received the same dose (13,700 IU) of vitamin D over 7 weeks, and a control group which received a daily Mongolian milk drink that did not contain vitamin D. The 5 different approaches to vitamin D supplementation were a daily tablet supplement, “seasonal supplementation” with tablet supplements given over 7 consecutive days, and 3 different daily milk drinks fortified with vitamin D: Mongolian milk, ultra-high temperature (UHT) milk, and a milk substitute. The primary endpoint of the trial was change in 25OHD concentrations. One of the 7 secondary endpoints was ARI. Parents were asked at baseline and study completion “Over the past 3 months, how many chest infections or ‘colds’ has your child had -counting only those infections that lasted for at least 24 hours with symptoms?” [4].

We obtained the publicly available dataset [7] and confirmed that the data for ARI matched the reported data [4]. We calculated the proportion of children with an ARI and the number of ARIs (0-6) by treatment group. Next, using recommended methodology, we estimated the ICC for ARI using the R ICCbin package, and the design effect for the study using the formula 1 + (M - 1) * ICC where M is the average cluster size [6]. Using the summary data on ARI and design effect, we calculated the risk reduction of ARI with vitamin D relative to the control group both for the different individual treatment groups and for the 5 vitamin D groups pooled together.

Next, we used the individual patient data to estimate the effect of vitamin D on the risk of having an ARI, and the number of ARIs, adjusting for the cluster trial design. The odds of an ARI were estimated using random-effects logistic regression using a random-intercept general linear model with a binomial distribution and logit link function. The effect on number of ARIs was estimated with random-effects negative binomial regression using the same approach but with a negative binomial distribution and a log link function to estimate rate ratios.

Finally, we updated the previous trial-level meta-analysis [2]. Data were extracted from the publication and random-effects meta-analyses run to replicate the published results. We then updated data for this study, adjusted for the cluster design, and re-ran the analyses. All analyses were performed using the R software package (R 3.5.1, 2019, R Foundation for Statistical Computing, Vienna, Austria)

## Results

### Analyses using summary data

Only limited basic demographics of the trial participants (gender, height, weight) are available in the public dataset [7], but full details have been reported by the investigators previously [3, 4, 7]. The number of participants, the number with ARI in the preceding 3 months at study end, and the number of individual episodes of ARI in the preceding 3 months at study end are shown in Table 1 and Table 2 by treatment group. Of note, the trial duration was only 7 weeks, so that some of the parental reported ARI in the preceding 3 months at study end may have occurred before the study began. The corresponding data match those published in the original article [4].

**Table 1:**
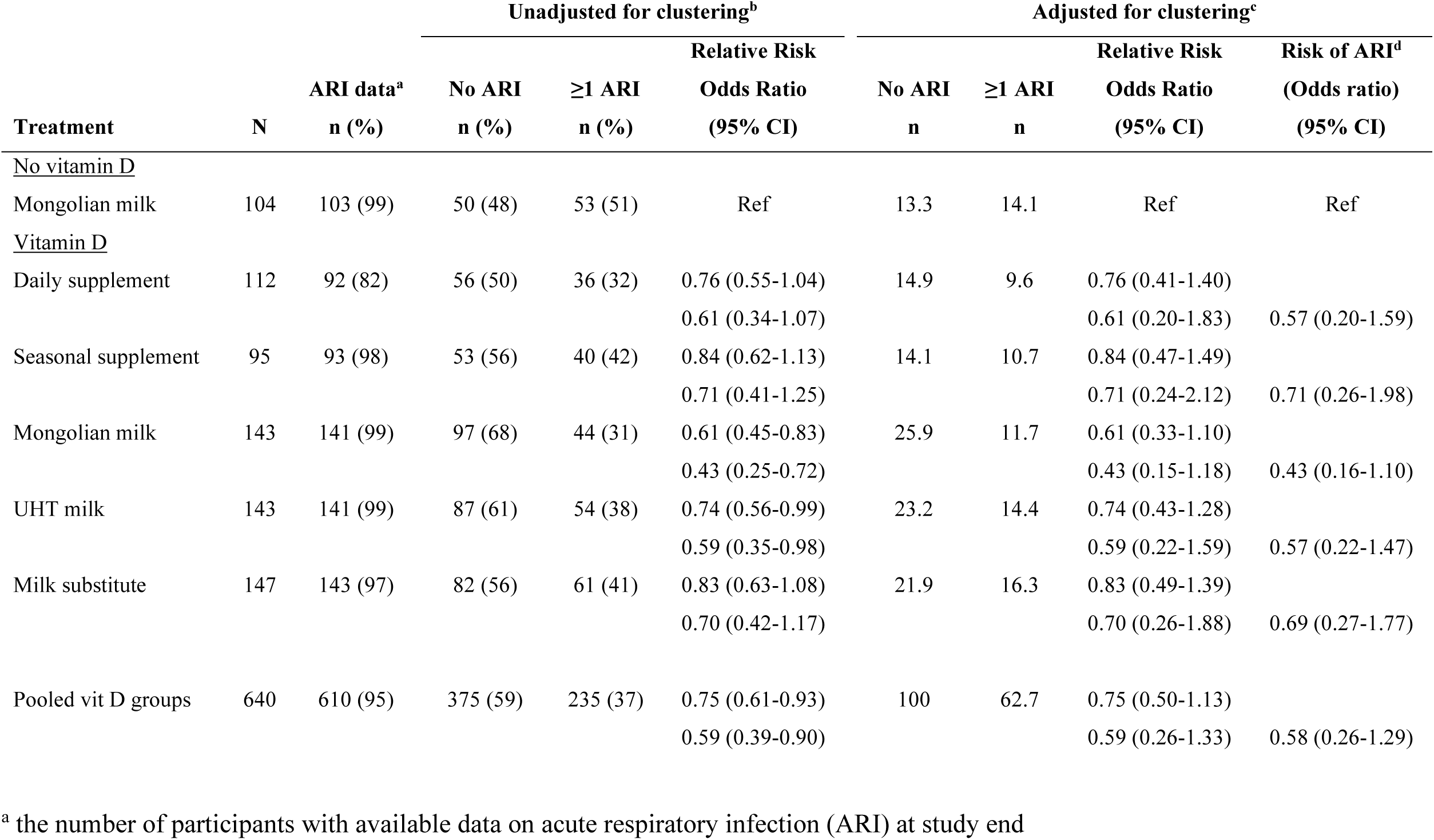
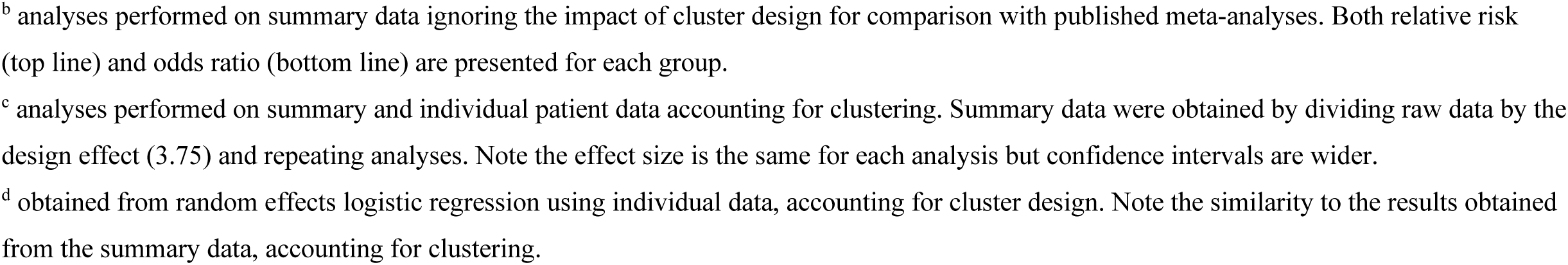
effect of vitamin D supplementation on the number of participants with ARI in the past 3 months.

| Treatment | N | ARI data<sup>a</sup><br>n (%) | Unadjusted for clustering<sup>b</sup> |  |  | Adjusted for clustering<sup>c</sup> |  |  |  |
| --- | --- | --- | --- | --- | --- | --- | --- | --- | --- |
|  |  |  | No ARI<br>n (%) | ≥1 ARI<br>n (%) | Relative Risk | No ARI<br>n | ≥1 ARI<br>n | Relative Risk | Risk of ARI<sup>d</sup> |
|  |  |  |  |  | Odds Ratio<br>(95% CI) |  |  | Odds Ratio<br>(95% CI) | (Odds ratio)<br>(95% CI) |
| <u>No vitamin D</u> |  |  |  |  |  |  |  |  |  |
| Mongolian milk | 104 | 103 (99) | 50 (48) | 53 (51) | Ref | 13.3 | 14.1 | Ref | Ref |
| <u>Vitamin D</u> |  |  |  |  |  |  |  |  |  |
| Daily supplement | 112 | 92 (82) | 56 (50) | 36 (32) | 0.76 (0.55-1.04) | 14.9 | 9.6 | 0.76 (0.41-1.40) |  |
|  |  |  |  |  | 0.61 (0.34-1.07) |  |  | 0.61 (0.20-1.83) | 0.57 (0.20-1.59) |
| Seasonal supplement | 95 | 93 (98) | 53 (56) | 40 (42) | 0.84 (0.62-1.13) | 14.1 | 10.7 | 0.84 (0.47-1.49) |  |
|  |  |  |  |  | 0.71 (0.41-1.25) |  |  | 0.71 (0.24-2.12) | 0.71 (0.26-1.98) |
| Mongolian milk | 143 | 141 (99) | 97 (68) | 44 (31) | 0.61 (0.45-0.83) | 25.9 | 11.7 | 0.61 (0.33-1.10) |  |
|  |  |  |  |  | 0.43 (0.25-0.72) |  |  | 0.43 (0.15-1.18) | 0.43 (0.16-1.10) |
| UHT milk | 143 | 141 (99) | 87 (61) | 54 (38) | 0.74 (0.56-0.99) | 23.2 | 14.4 | 0.74 (0.43-1.28) |  |
|  |  |  |  |  | 0.59 (0.35-0.98) |  |  | 0.59 (0.22-1.59) | 0.57 (0.22-1.47) |
| Milk substitute | 147 | 143 (97) | 82 (56) | 61 (41) | 0.83 (0.63-1.08) | 21.9 | 16.3 | 0.83 (0.49-1.39) |  |
|  |  |  |  |  | 0.70 (0.42-1.17) |  |  | 0.70 (0.26-1.88) | 0.69 (0.27-1.77) |
| Pooled vit D groups | 640 | 610 (95) | 375 (59) | 235 (37) | 0.75 (0.61-0.93) | 100 | 62.7 | 0.75 (0.50-1.13) |  |
|  |  |  |  |  | 0.59 (0.39-0.90) |  |  | 0.59 (0.26-1.33) | 0.58 (0.26-1.29) |

<sup>a</sup> the number of participants with available data on acute respiratory infection (ARI) at study end
<sup>b</sup> analyses performed on summary data ignoring the impact of cluster design for comparison with published meta-analyses. Both relative risk (top line) and odds ratio (bottom line) are presented for each group.
<sup>c</sup> analyses performed on summary and individual patient data accounting for clustering. Summary data were obtained by dividing raw data by the design effect (3.75) and repeating analyses. Note the effect size is the same for each analysis but confidence intervals are wider.
<sup>d</sup> obtained from random effects logistic regression using individual data, accounting for cluster design. Note the similarity to the results obtained from the summary data, accounting for clustering.

**Table 2:** effect of vitamin D supplementation on the number of ARI in the past 3 months.

| Treatment | N | ARI data<sup>a</sup><br>n (%) | Number of ARI [n (%)] |  |  |  |  |  |  | Rate Ratio<br>(95%CI)<sup>b</sup> |
| --- | --- | --- | --- | --- | --- | --- | --- | --- | --- | --- |
|  |  |  | 0 | 1 | 2 | 3 | 4 | 5 | 6 |  |
| <u>No vitamin D</u> |  |  |  |  |  |  |  |  |  |  |
| Mongolian milk | 104 | 103 (99) | 50 (48) | 32 (31) | 14 (13) | 6 (6) | 1 (1) | 0 (0) | 0 (0) | Ref |
| <u>Vitamin D</u> |  |  |  |  |  |  |  |  |  |  |
| Daily supplement | 112 | 92 (82) | 56 (50) | 24 (21) | 11 (10) | 0 (0) | 1 (1) | 0 (0) | 0 (0) | 0.68 (0.38-1.24) |
| Seasonal supplement | 95 | 93 (98) | 53 (56) | 31 (33) | 7 (7) | 1 (1) | 1 (1) | 0 (0) | 0 (0) | 0.72 (0.40-1.30) |
| Mongolian milk<sup>c</sup> | 143 | 141 (99) | 97 (68) | 31 (22) | 10 (7) | 1 (1) | 1 (1) | 0 (0) | 1 (1) | 0.58 (0.34-1.01) |
| UHT milk | 143 | 141 (99) | 87 (61) | 35 (24) | 14 (10) | 4 (3) | 0 (0) | 1 (1) | 0 (0) | 0.73 (0.43-1.26) |
| Milk substitute | 147 | 143 (97) | 82 (56) | 41 (28) | 13 (9) | 4 (3) | 1 (1) | 2 (1) | 0 (0) | 0.80 (0.47-1.37) |
| Pooled vit D groups | 640 | 610 (95) | 375 (59) | 162 (25) | 55 (9) | 10 (2) | 4 (1) | 3 (0) | 1 (0) | 0.70 (0.44-1.12) |

<sup>a</sup> the number of participants with available data on acute respiratory infection (ARI) at study end
<sup>b</sup> obtained from random effects negative binomial regression accounting for cluster design
<sup>c</sup> data included in original trial report

There were 744 participants in the trial, randomized to 6 treatment groups, in 21 different clusters. The mean number of participants per cluster was 35.4. The ICC for ARI at study end was 0.08 (95% CI 0.02-0.14). Therefore, the design effect is 3.75, indicating that summary statistics of data unadjusted for clustering should be decreased by 76% to account for the cluster design [6]. Table 1 shows the effect of this adjustment on the effect of vitamin D supplementation on the proportions of children with ARI: the relative risk and odds ratios remain unchanged, but the confidence intervals are much wider because the sample size is effectively much smaller. After taking account of clustering, there is no statistically significant effect of vitamin D supplementation on children experiencing ARI in any treatment group, or when the vitamin D groups are pooled and analysed together.

### Analyses using individual data

The effect of vitamin D supplementation on the proportions of individuals with ARI adjusted for the cluster design is shown in Table 1, and the effect of vitamin D on the number of episodes of ARI adjusted for the cluster design in Table 2. In each analysis, there is no statistically significant effect of vitamin D supplementation in any treatment group, or when the vitamin D groups are pooled and analysed together.

Carmago reported that, when comparing the Mongolian milk with vitamin D vs Mongolian milk without vitamin D groups, vitamin D reduced the number of ARI episodes: n= 244, rate ratio 0.52 (0.31-0.89) [4]. We restricted our analyses to the same two treatment groups, using number of ARI as the endpoint, and a random intercept negative binomial regression model. Using R (glmer function), the results were n=244, rate ratio 0.58 (0.34-0.99). The original analyses were conducted using Stata. We wondered whether the small difference between the results was due to difference in statistics programs. We repeated the analysis in Stata (xtnbreg function) and replicated the original results. We then repeated the analysis in SAS (genmod procedure) and obtained similar results to those from R: 0.58 (0.34-0.97). We are unsure of the reasons underlying these small differences, but speculate that they may be due to differences in handling of the dispersion parameter between the programs.

### Re-analysis of trial-level meta-analysis

We extracted data from the publication and replicated the published results: odds ratio (OR) 0.92 (0.86-0.99), I^2^= 36%; relative risk (RR) 0.98 (0.95-1.00) [2]. We then re-analysed these data firstly using the summary statistics from Table 1 for only the two trial arms of Carmago 2012 that were included in the original meta-analysis. The trial results changed from 44/141 vs 53/103, OR 0.43 (0.25-0.72) to 11.7/37.6 vs 14.1/27.4, OR 0.43 (0.16-1.10), and the pooled meta-analysis results to OR 0.94 (0.88-1.01), I^2^= 27%, P=0.07; RR 0.98 (0.96-1.00), P=0.10.

We repeated these analyses using the summary statistics from Table 1 for the comparison of the pooled five vitamin D groups with the control group (62.7/162.7 vs 14.1/27.4, OR 0.59, 0.26,1.33, Table 1) instead of the existing Camargo 2012 data used in the original meta-analysis. Using these results, the pooled result for the meta-analysis is: 37 trials, OR 0.94 (0.88-1.01), I^2^ =25%, P=0.07; RR 0.98 (0.96-1.00), P=0.10 (Figure 1). While OR was used in the original analyses [2], as the incidence of ARI was not uncommon (62%), the OR is likely to overestimate the RR. We therefore have presented calculated RR throughout.

**Figure 1:**
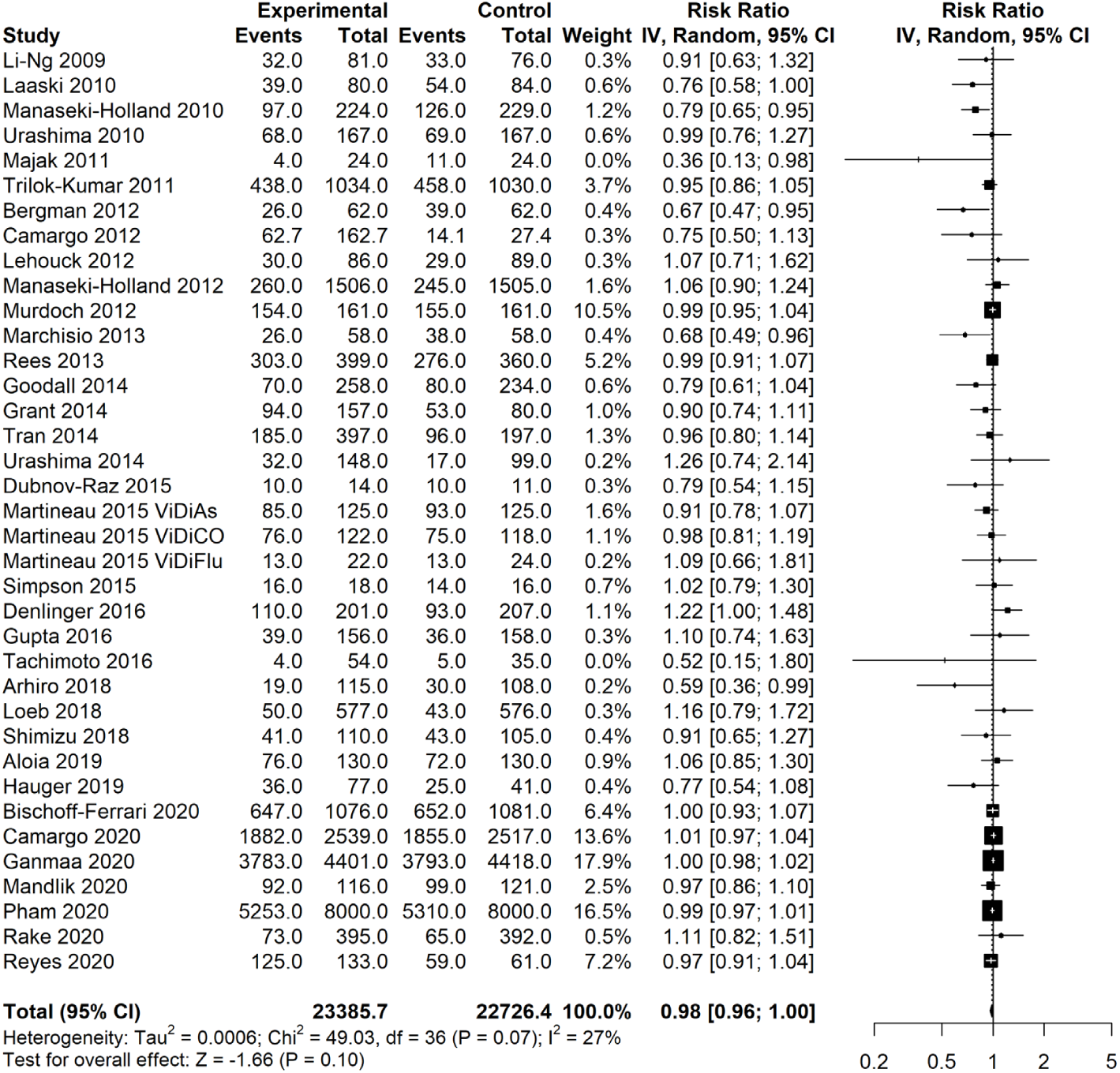
meta-analyses from Jolliffe and colleagues [2] updated with data for Camargo 2012 that includes all five vitamin D supplementation arms and is correctly adjusted for the cluster-randomised design. If only the two trial arms used in the original meta-analyses are used for Camargo 2012, the pooled result and p-value are unchanged but I^2^ = 28%.

## Discussion

Here we present additional data to those presented previously in Carmago 2012 [4]. The current data provide additional information, in the form of the ICC (0.08) and design effect (3.75), that allows researchers to correctly use the original reported summary data in meta-analyses. We also provide estimates of the effect of vitamin D on ARI from four previously unpublished groups of the trial, which in turn can be used in future meta-analyses. These newly reported results show that vitamin D supplementation had no statistically significant effect on the proportion of individuals with an ARI or on the number of ARIs in this trial. When these data are used to update the latest trial-level meta-analysis [2], the pooled results for vitamin D, either when the Blue Sky trial data are restricted to the two vitamin D arms used in the original meta-analysis or when all vitamin D arms are incorporated, are neutral with the confidence intervals including unity. The earlier individual patient data meta-analysis [1], for which the data are not currently publicly available, should also be updated taking into account the cluster design of this trial.

As of February 2022, the two meta-analyses [1, 2] have been cited 48 and 881 times respectively, in Scopus. Searching for “Covid” within the 905 citing documents returned 526 articles, many citing these two meta-analyses as evidence that vitamin D supplements may reduce the risk of Covid-19 or mitigate the illness. The updated pooled trial-level meta-analysis results do not support suggestions that vitamin D supplementation should be prescribed to prevent Covid-19.

## Data Availability

Data sharing: All data are publicly available from the primary publications listed in the manuscript.

## Acknowledgement

Although not involved with the current analyses, we wish to acknowledge Associate Janet Rich-Edwards, Harvard T.H. Chan School of Public Health, and her colleagues who undertook the trial and made the trial dataset publicly available, and Associate Professor Rich-Edwards for supporting further analyses of this dataset.

